# Development and Validation of an Artificial Intelligence Predictive Model to Accelerate Antibiotic Therapy for Critical Ill Children with Sepsis in the Pediatric ED with Pediatric ICU Disposition

**DOI:** 10.1101/2025.03.25.25324127

**Authors:** Kathleen Cao, Nikolay Braykov, Andrea McCarter, Swaminathan Kandaswamy, Evan W. Orenstein, Edwin Ray, Rebekah Carter, Mary Beth Gleeson, Srikant Iyer, Naveen Muthu, Mark V. Mai

## Abstract

**Importance:** Pediatric sepsis accounts for over 72,000 US hospitalizations annually with significant mortality and morbidity. Many pediatric hospitals struggle to promptly identify and treat sepsis. This study demonstrates the feasibility of a multi-tiered artificial intelligence (AI) to enhance sepsis clinical decision-making within a complex emergency department (ED) workflow.

**Objectives:** To develop and validate a local AI model predicting critical sepsis among ED patients who received a fluid bolus and a disposition to the Pediatric Intensive Care Unit (PICU) but had not yet received antibiotics.

**Design:** Retrospective observational cross-section study

**Setting:** Urban, quaternary-care, academic healthcare system

**Patients:** Pediatric ED patients

**Interventions:** None

**Measures and Main Results:** The “Sepsis on ED to PICU Disposition” (SEPD) model aimed to predict critical sepsis within 72 hours of PICU disposition using a dataset totaling 5,534 patient encounters for model training and testing. During silent implementation, 1,058 encounters were used for validation. The SEPD model outperformed a vendor-developed sepsis model with an AUROC of 81.8%, compared to 57.5%. The model also demonstrated better precision-recall performance, showing more balanced identification of true positives. During silent implementation, the SEPD model maintained similar sensitivity (85.29%) and specificity (60.45%) to those observed during model testing.

**Conclusion:** The SEPD model improved detection of critical sepsis among high-risk pediatric ED patients with a known PICU disposition, outperforming a vendor-developed sepsis model. Within a complex ED workflow, this model may facilitate timely sepsis identification and treatment in critically ill patients, who may have been missed during earlier stages of their ED course.

## INTRODUCTION

Pediatric sepsis accounts for over 72,000 hospitalizations in the United States and is associated with significant mortality and morbidity.^1–3^ Current Surviving Sepsis guidelines recommend that antibiotics be delivered within one hour for septic shock and within three hours for severe sepsis identification to reduce the increased risk of mortality.^4–6^ Despite this recommendation, many pediatric hospitals struggle with timely identification and management of sepsis. Early diagnosis is key in reducing the risk of mortality and morbidity associated with sepsis. Predictive models using artificial intelligence (AI) have been touted as potential solutions to improving situational awareness of patients at risk of becoming septic.^7–11^

Our electronic health record (EHR) vendor, Epic Systems© (Epic Systems, Verona, WI) developed a pediatric sepsis model in the emergency room (ED) in collaboration with Nationwide Children’s Hospital that incorporates exam findings, vital sign values, and presence of high-risk conditions to generate a risk score of sepsis among all comers to the pediatric ED.^12^ We previously described our experience implementing this vendor-developed model.^13^ Briefly, the vendor-developed model was silently implemented in our health system without provider-facing alerts beginning 2019 to evaluate its predictive performance against sepsis, as defined by the Improving Pediatric Sepsis Outcomes (IPSO) sepsis criteria.^14,15^ Following silent implementation and triggering criteria optimization, the model was implemented as an interruptive nurse-facing alert that screens all ED patients for sepsis, prompting nurse-physician huddles to rapidly assess the need for fluid boluses and antibiotics.

While the model improved the time to first fluid bolus, time to first antibiotics did not initially improve. On further investigation, we found that many of the ED critical sepsis patients with prolonged time to first antibiotic did not receive the antibiotic until arrival in the Pediatric Intensive Care Unit (PICU). Additionally, we found that while among all comers to the ED, only ∼1/300 met IPSO sepsis criteria, the baseline prevalence was much higher among children who had received a fluid bolus but had not yet received a sepsis antibiotic at the time of disposition to the PICU (prevalence 8.0% for any IPSO sepsis, 3.9% for IPSO critical sepsis). Identifying this subpopulation at high risk for sepsis facilitated the development of a targeted predictive model to improve sepsis management within the ED workflow.

In this study, we aimed to develop and validate a custom AI model predicting IPSO critical sepsis among ED patients who received a fluid bolus but had yet to receive antibiotics at the time of ICU disposition. In a companion paper, we describe the usability testing and implementation outcomes of clinical decision support based on the model developed here.

## METHODS

### Study Design and Population

This study describes an observational cross-sectional study involving the development (starting November 2022) and validation (starting June 2023) through silent implementation of a single-center custom AI prediction model entitled “Sepsis on ED to PICU Disposition” (SEPD). This model was deployed in the ED at a large quaternary pediatric health system in the Southeastern US across 2 campuses, each with over 165,000 ED visits per year. Given that the development of this model occurred as part of a quality improvement initiative, the protocol for this retrospective analysis of model performance was reviewed and deemed not human subjects research by the Children’s Healthcare of Atlanta Institutional Review Board. No funding source or conflicts of interest were identified for this study.

A retrospective patient cohort was developed through query of existing data in our electronic health record (Epic© Systems, Verona, WI) involving all pediatric ED visits from August 2020 to September 2022who received a fluid bolus in the ED, were dispositioned to the PICU, but did not have sepsis antibiotics administered in the ED as of the time of disposition entry by the ED physician. This retrospective cohort was randomly split to 80% for model training and 20% for testing. Evaluation of the predictive model followed the transparent reporting of a multivariable prediction model for individual prognosis or diagnosis (TRIPOD) recommendations.^16,17^

### Model Development

The primary outcome label was critical sepsis, defined using the Children’s Hospital Association’s Improving Sepsis Outcomes (IPSO) collaborative definition for critical sepsis.^14^ To meet the IPSO critical sepsis definition, patients must receive (1) intravenous antibiotics and (2) at least three boluses or two or more boluses and a vasoactive agent. These two criteria must have occurred within 6 hours of each other, and a blood culture must also have been collected within 72 hours of the antibiotic and bolus criteria. The model was trained to predict critical sepsis within 72 hours from patient disposition. Patients with disposition outside of the PICU were excluded.

Candidate features were developed through review of literature and a multidisciplinary team consisting of physicians, nurses, informaticists, and data scientists. Initial features for inclusion in the model consisted of demographics, financial class, location (campus and trauma room), lab values (C-reactive protein (CRP), Procalcitonin, and Neutrophils), Epic sepsis model score, and number of high-risk conditions (including technology dependent, static encephalopathy, malignancy, sickle cell disease, history of organ transplant, immunosuppression, chemotherapy treatment plan, and presence of a central venous access device (CVAD)). Statistical significance of univariable associations between candidate variables and IPSO critical sepsis was calculated using the Wald chi-square statistic, which sorts the variables by their coefficient value divided by the standard error (Figure 1). Supplemental Table 1 represents all model predictor variables and criteria considered with details.

**Figure 1.**
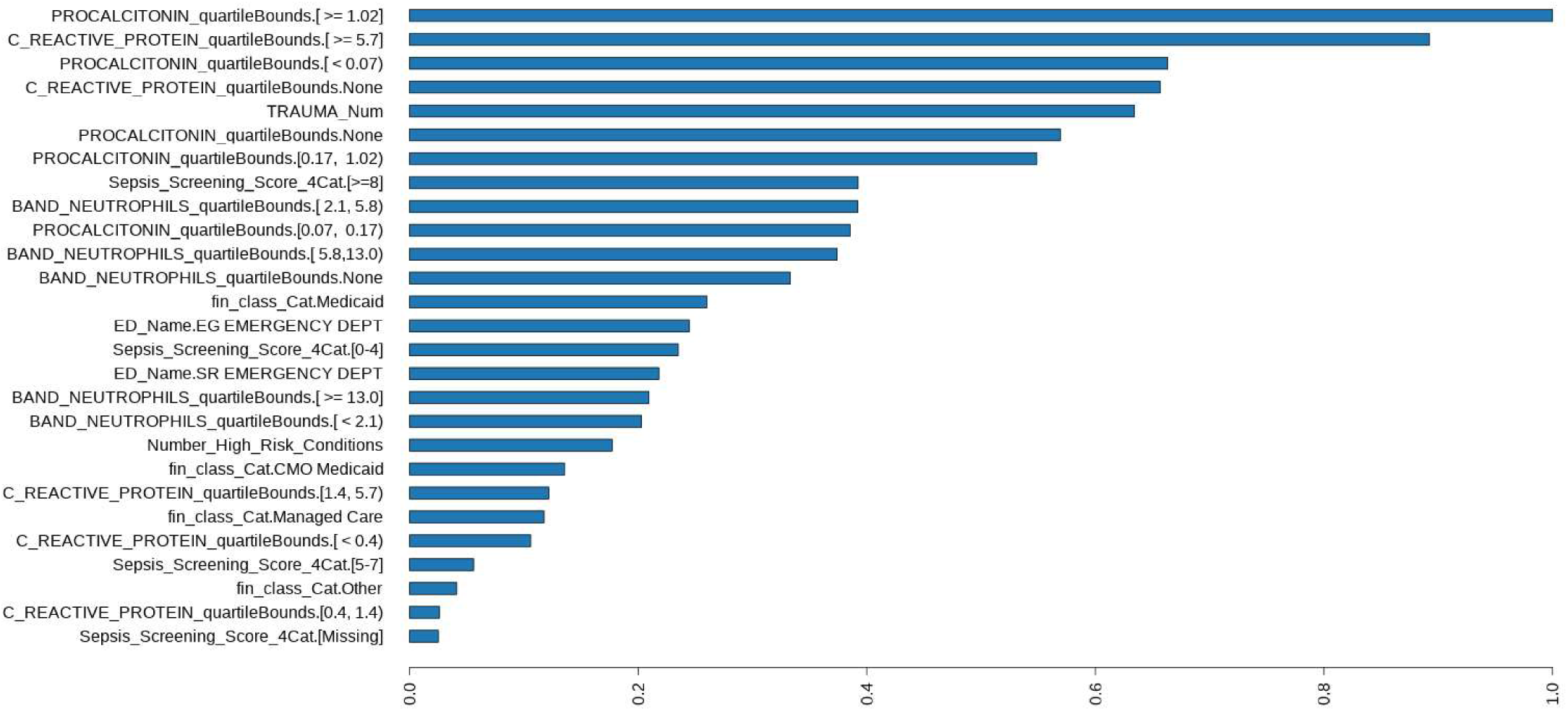
Top candidate variables were ranked by variable importance. Variable importance was ranked by statistical significance via the Wald chi-square statistic. Top variables included laboratory markers associated with inflammation (e.g. procalcitonin, c-reactive protein), as well as demographic (e.g. Medicaid, high-risk conditions) and workflow variables (e.g. trauma room).

Models were developed using generalized linear models (GLMs) and random forests to predict the primary outcome. A model evaluation dashboard was created to compare iterations of 64 different algorithms and feature sets adjusting for considered predictor variables to select the best model by overall performance benchmark metric using area under the receiver operating characteristic curve (AUROC) and area under the precision recall curve (AUPRC) on the test data set. Performance metrics were evaluated for each model with ten-fold cross-validation.

Missing values for all lab values were coded as “None”, for financial class as “Other”, for number of high-risk conditions as “0”, for trauma room as “1” (not in trauma room), and for EPIC sepsis scores as “Missing”. Logistic ridge regression with regularization (30 lambda values evaluated), and outcome balancing with ROSE (Random Over-Sampling Examples) were found to provide the best metrics. Final variable importance (Figure 1) reflects the magnitude of the coefficient value divided by the standard error for each variable, which indicates the level of the significance of the relationship between the variable and the outcome.

The Epic pediatric ED sepsis score was used as a baseline comparator for model performance. High performing models were reviewed by clinical experts for face validity of key features and by EHR implementation engineers for effort estimation of translating the model into the production system.

### Silent Implementation and Evaluation

Features in the final selected model were built as reporting workbench columns in Epic Systems. The final candidate model was represented using predictive markup model language (PMML) and implemented into the live production system on June 7^th^, 2023, and scores were filed every 15 minutes. However, at that time no scores or alerts were shown to clinical users. The models ran on all patients who had received a fluid bolus, had not received antibiotics, had not had a sepsis huddle completed, and were not ED boarders (admitted patients waiting in the ED > 2 hours for an inpatient bed). Model performance was evaluated by selecting the sepsis score filed closest to time of PICU disposition for a particular encounter (ex: within 30 minutes, before or after) and compared against the label of eventual inclusion in IPSO cohort in next 72 hours.

Sensitivity of 75% and positive predictive value of 7.5% were set as target benchmarks based on prior qualitative work.^13^ Initial scores were filed every 15 minutes on the target population and changed on July 8^th^, 2023, to file every hour due to performance concerns. The threshold of 0.37 was set based on an F2 score to balance precision and recall in the setting of increased patient risk with false negatives.

## RESULTS

### Study Population

A total of 5,534 patient encounters between August 2020 to September 2022 were used for development and testing of this model. Silent implementation from June 7th, 2023, to December 7th, 2023, consisted of a total of 1,058 patient encounters. Demographic description of the entire cohort during training, testing, and silent implementation consisted of gender, race, ethnicity, financial class, and ≥1 complex chronic condition (Table 1). Age at time of arrival was evaluated separately in test models as a possible predictor. Demographics of the patients in the training, testing and silent implementation were similar (Table 1). During model training, the characteristic of ≥1 Complex Chronic Condition was statistically significant (p <0.001) for patients with critical sepsis (n = 61, 36.1%) vs. no critical sepsis (n = 869, 20.4%). This significance was not replicated in testing or silent implementation cohorts.

**Table 1:** Demographics of Model Development Training, Test, and Prospective Silent Implementation Cohorts^a^.

|  |  | Development - Training |  |  |  | Development - Test |  |  |  | Prospective Silent Implementation |  |  |  |
| --- | --- | --- | --- | --- | --- | --- | --- | --- | --- | --- | --- | --- | --- |
| Characteristic |  | All<br>(n = 4,427) | No Critical<br>Sepsis<br>(n = 4258) | Critical<br>Sepsis<br>(n = 169) | P value | All<br>(n = 1,107) | No Critical<br>Sepsis<br>(n = 1062) | Critical<br>Sepsis<br>(n = 45) | P value | All<br>(n = 1058) | No Critical<br>Sepsis<br>(n = 1024) | Critical<br>Sepsis<br>(n = 34) | P value |
| Gender |  |  |  |  | 0.953 |  |  |  | 0.023 |  |  |  | 0.203 |
|  | Female | 1968<br>(44.5%) | 1892<br>(44.4%) | 76 (45.0%) |  | 494 (44.6%) | 466 (43.9%) | 28 (62.2%) |  | 463 (43.8%) | 444 (43.4%) | 19 (55.9%) |  |
|  | Male | 2459<br>(55.5%) | 2366<br>(55.6%) | 93 (55.0%) |  | 613 (55.4%) | 596 (56.1%) | 17 (37.8%) |  | 595 (56.2%) | 580 (56.6%) | 15 (44.1%) |  |
| Race |  |  |  |  | 0.772 |  |  |  | 0.857 |  |  |  | 0.124 |
|  | Black or<br>African<br>American | 1738<br>(39.3%) | 1668<br>(39.2%) | 70 (41.4%) |  | 424 (38.3%) | 405 (38.1%) | 19 (42.2%) |  | 411 (38.8%) | 397 (38.8%) | 14 (41.2%) |  |
|  | White | 1978<br>(44.7%) | 1907<br>(44.8%) | 71 (42.0%) |  | 495 (44.7%) | 476 (44.8%) | 19 (42.2%) |  | 499 (47.2%) | 487 (47.7%) | 12 (35.3%) |  |
|  | Other | 711 (16.1%) | 683 (16.0%) | 28 (16.6%) |  | 188 (17.0%) | 181 (17.0%) | 7 (15.6%) |  | 110 (10.4%) | 103 (10.0%) | 7 (20.6%) |  |
|  | Declined | - | - | - |  | - | - | - |  | 38 (3.59%) | 37 (3.61%) | 1 (2.94%) |  |
| Ethnicity |  |  |  |  | 0.734 |  |  |  | 0.558 |  |  |  | 0.212 |
|  | Hispanic or<br>Latino | 790 (17.8%) | 762 (17.9%) | 28 (16.6%) |  | 196 (17.7%) | 190 (17.9%) | 6 (13.3%) |  | 179 (16.9%) | 177 (17.3%) | 2 (5.88%) |  |
|  | Non-Hispanic<br>or Unknown | 3637<br>(82.2%) | 3496<br>(82.1%) | 141 (83.4%) |  | 911 (82.3%) | 872 (82.1%) | 39 (86.7%) |  | 879 (83.1%) | 847 (82.7%) | 32 (94.1%) |  |
| Insurance |  |  |  |  | 0.575 |  |  |  | 0.814 |  |  |  | 0.279 |
|  | Public | 2849<br>(64.4%) | 2736<br>(64.3%) | 113 (66.8%) |  | 721 (65.1%) | 689 (64.9%) | 32 (71.1%) |  | 663 (62.7%) | 642 (62.7%) | 21 (61.8%) |  |
|  | Private | 1492<br>(33.7%) | 1439<br>(33.8%) | 53 (31.4%) |  | 369 (33.4%) | 356 (33.5%) | 13 (28.9%) |  | 368<br>(34.8%) | 355 (34.7%) | 13 (38.2%) |  |
|  | Self-Pay | 5 (0.11%) | 5 (0.12%) | 0 (0.00%) |  | 2 (0.18%) | 2 (0.19%) | 0 (0.00%) |  | - | - | - |  |
|  | Unknown | 81 (1.83%) | 78 (1.83%) | 3 (1.78%) |  | 15 (1.36%) | 15 (1.41%) | 0 (0.00%) |  | 27<br>(2.55%) | 27 (2.64%) | 0 (0.00%) |  |
| Age at Time of<br>Arrival |  | 4.25<br>[1.25;11.7] | 4.25<br>[1.25;11.7] | 5.25<br>[1.00;13.5] | 0.496 | 4.33<br>[1.12;12.1] | 4.25<br>[1.17;12.0] | 4.83<br>[0.17;13.2] | 0.430 | 3.17<br>[0.60;9.31] | 3.17<br>[0.67;9.33] | 3.54<br>[0.17;6.94] | 0.870 |

<sup>a</sup>ED patients treated with a fluid bolus and no antibiotics prior to entry of disposition to the Pediatric ICU.

### Model Performance

During model training, the SEPD model performed better than the existing Epic Sepsis score model using the AUROC curve which summarizes the trade-off between sensitivity (true positive rate or recall) and specificity (true negative rate) of a classifier. The SEPD model had a higher AUROC of 81.8% compared to 57.5% (Figure 2a and 2b). The SEPD model also had a higher AUPRC compared to the existing sepsis model (Figure 2c and 2d).

**Figure 2:**
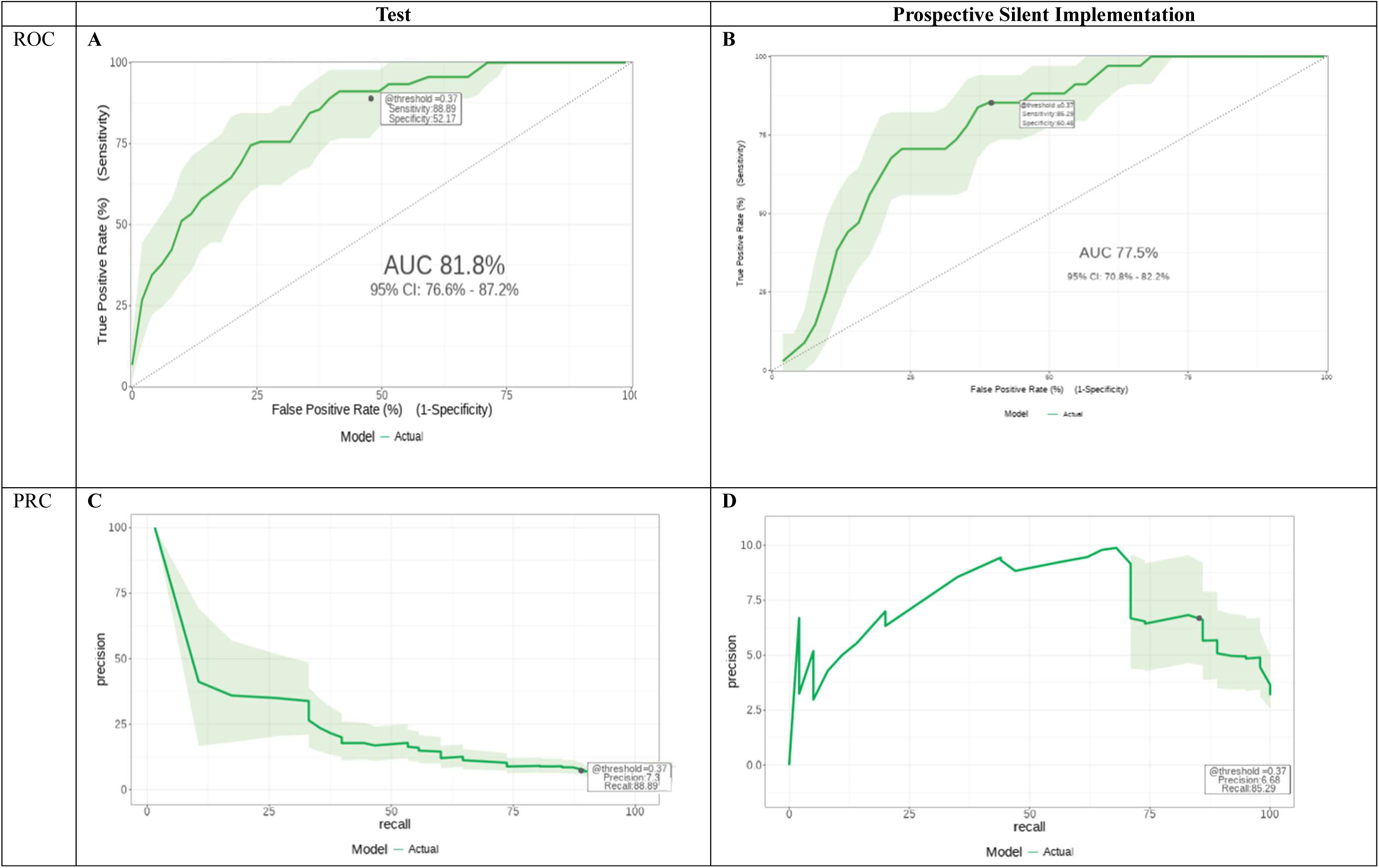
Model performance of the SEPD model was similar during model testing and silent implementation. Panels A and B show the ROC curves for the test dataset and prospective silent implementation, respectively, with AUCs of 81.8% (95% CI: 76.0%– 87.2%) and 77.5% (95% CI: 70.8%–82.2%). Panels C and D display the corresponding PRC curves, highlighting precision versus recall across thresholds for the test dataset and prospective silent implementation. Note Shaded regions represent 95% confidence intervals.

The final model consisted of the variables identified in Table 2, sorted by importance. The most important variables were procalcitonin ≥1.02, CRP ≥5.7, and if the patient had been in the trauma room, where the most acute patients are seen, regardless of whether trauma was present on arrival. The variable “Number of High-risk conditions” was found to provide more information for the model and was included in the final model. The initial model included disposition requirement but ended up firing too many false positives and was revitalized to closest score to disposition (1 hour within disposition). Model code and formula can be found in Supplemental Table 1 and Supplemental Figure 1.

**Table 2:**
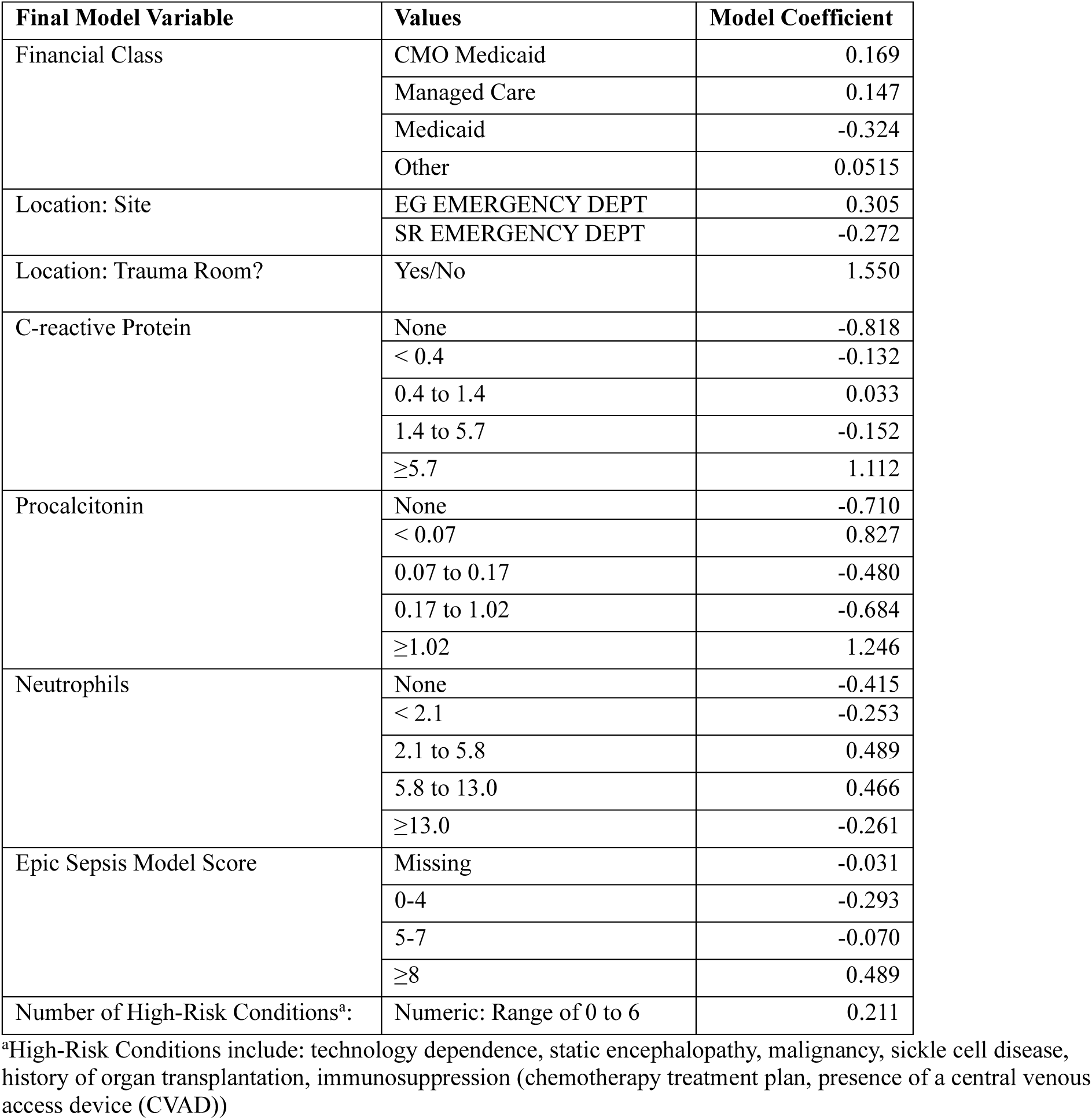
Final Model Variables, Values, and Coefficients.

| Final Model Variable | Values | Model Coefficient |
| --- | --- | --- |
| Financial Class | CMO Medicaid | 0.169 |
|  | Managed Care | 0.147 |
|  | Medicaid | -0.324 |
|  | Other | 0.0515 |
| Location: Site | EG EMERGENCY DEPT | 0.305 |
|  | SR EMERGENCY DEPT | -0.272 |
| Location: Trauma Room? | Yes/No | 1.550 |
| C-reactive Protein | None | -0.818 |
|  | < 0.4 | -0.132 |
|  | 0.4 to 1.4 | 0.033 |
|  | 1.4 to 5.7 | -0.152 |
|  | ≥5.7 | 1.112 |
| Procalcitonin | None | -0.710 |
|  | < 0.07 | 0.827 |
|  | 0.07 to 0.17 | -0.480 |
|  | 0.17 to 1.02 | -0.684 |
|  | ≥1.02 | 1.246 |
| Neutrophils | None | -0.415 |
|  | < 2.1 | -0.253 |
|  | 2.1 to 5.8 | 0.489 |
|  | 5.8 to 13.0 | 0.466 |
|  | ≥13.0 | -0.261 |
| Epic Sepsis Model Score | Missing | -0.031 |
|  | 0-4 | -0.293 |
|  | 5-7 | -0.070 |
|  | ≥8 | 0.489 |
| Number of High-Risk Conditions<sup>a</sup>: | Numeric: Range of 0 to 6 | 0.211 |

<sup>a</sup>High-Risk Conditions include: technology dependence, static encephalopathy, malignancy, sickle cell disease, history of organ transplantation, immunosuppression (chemotherapy treatment plan, presence of a central venous access device (CVAD))

**Table 3:** Model Performance Metrics of the SEPD Predictive Model During Training, Testing, and Silent Implementation.

| Model | Derivation - Training | Derivation - Test | Prospective Silent Implementation |
| --- | --- | --- | --- |
| Patient Total | 4427 | 1107 | 1058 |
| Area under the ROC Curve | 0.802 | 0.818 | 0.775 |
| Performance at threshold 0.37* |  |  |  |
| ▪ Sensitivity (%) | 88.76 | 88.89 | 85.29 |
| ▪ Specificity (%) | 53.92 | 52.17 | 60.45 |
| ▪ PPV (%) | 7.10 | 7.30 | 6.68 |
| ▪ NPV (%) | 99.2 | 99.1 | 99.2 |

\*Selected cutoff based off the F2 score, prioritizing sensitivity over specificity with goal minimum sensitivity 75% and PPV of 7.5%

Model performance was compared during the training, testing and silent implementation period with selected cutoff based on F2 score of 0.37 (Table 3). Performance between the test population and background prospective silent implementation were similar with AUROCs of 81.8% and 77.5% respectively. Additionally, testing versus silent implementation sensitivity (88.89% vs. 85.29%) and PPV (7.30% vs. 6.68%) were overall similar with the prospective silent implementation showing better specificity compared to the test performance (60.45% vs 52.17%). We did not find significant differences in model sensitivity and positive predictive value by patient race, sex, ethnicity, primary language, insurance status or age (Supplemental Tables 2 and 3)

## DISCUSSION

As part of a quality improvement initiative to improve sepsis outcomes in the ED, we developed and internally validated a custom AI model called SEPD predicting IPSO critical sepsis among ED patients who received a fluid bolus with an ICU disposition but had yet to receive antibiotics. Key model variables included inflammatory markers, a measure of patient fragility, and a measure of patient acuity. Performance of the SEPD model was consistent during model evaluation through silent implementation and maintained better predictive performance compared to a vendor-developed model during a 6-month period of silent implementation (Figure 2). The vendor-developed sepsis model is currently deployed as a screening step earlier in the ED sepsis workflow, while the SEPD model is deployed as a second stage model at the time of patient disposition from the ED.

To our knowledge, this is the first description of a single clinical workflow incorporating multiple prediction models at different clinical decision-making steps in the same clinical setting. In concert, the vendor-developed ED sepsis model plus the custom developed SEPD model help stratify and identify patients at high risk of developing sepsis. Other studies have described implementations of single alerts to address timely delivery of sepsis fluids and antibiotics.^18–21^ At our institution, use of the vendor-developed model for nurse-led sepsis huddles improved time to fluid boluses, but not antibiotics. While the vendor-developed model helped to improve situational awareness for patients at risk, the challenge with such a model is that the overall prevalence of sepsis in all patients seen in the ED is exceedingly rare (0.003%), creating a difficult tradeoff between adequate sensitivity and number needed to alert. During the process of reviewing opportunities for improvement of the AI-augmented workflow, we found that by selecting a later point in the workflow, when ICU disposition had already been decided and a fluid bolus had already been administered, we could enrich for a population more likely to develop sepsis (IPSO sepsis 8.0%, IPSO critical sepsis 3.9%). As described in the companion paper, implementation of this model resulted in a significant decrease in time to antibiotic administration across both campuses. Such a framework – a screening model with a downstream model enriched using restricting criteria – may be a helpful approach in low-prevalence, high-risk conditions in complex clinical workflows.

Limitations of our study include model development and validation at a single institution which may affect generalizability based on differences in populations at other centers. Specifically, some model features such as the patient being admitted to a trauma room may be highly variable in informing model predictions as institutions have variation in their triage protocols. This model also excludes patients who do not have disposition to the PICU from the ED which could result in missed opportunities for early initiation of antibiotics in pediatric patients in the ED designated for general inpatient admission. Additionally, development of the SEPD model only describes progress up to the silent implementation phase but does not include real-time implementation in a high acuity unit to assess feasibility and effectiveness of the model in managing pediatric patients in the ED. A companion paper will address this issue and describe usability testing and evaluate the impact of implementation of clinical decision support in the ED using the SEPD model.

## CONCLUSION

As part of a multi-tiered AI approach to sepsis detection in the pediatric ED, a locally developed model targeting critical sepsis among high-risk patients bound for the PICU demonstrated improved predictive performance compared to an existing vendor-developed model. While limited to a single healthcare system across two hospitals, this model highlights the potential of AI to improve the timeliness of sepsis treatment at multiple points along a clinical workflow. This model may help clinicians identify patients at particularly high risk of developing sepsis missed by the initial screen, potentially reducing delays in treatment and improving outcomes for critically ill children.

## Data Availability

Data sharing is not applicable to this article as no new datasets were generated or analyzed during the quality improvement project. This study used existing institutional data that cannot be shared publicly due to confidentiality agreements.

---

**Supplemental Table 1:**
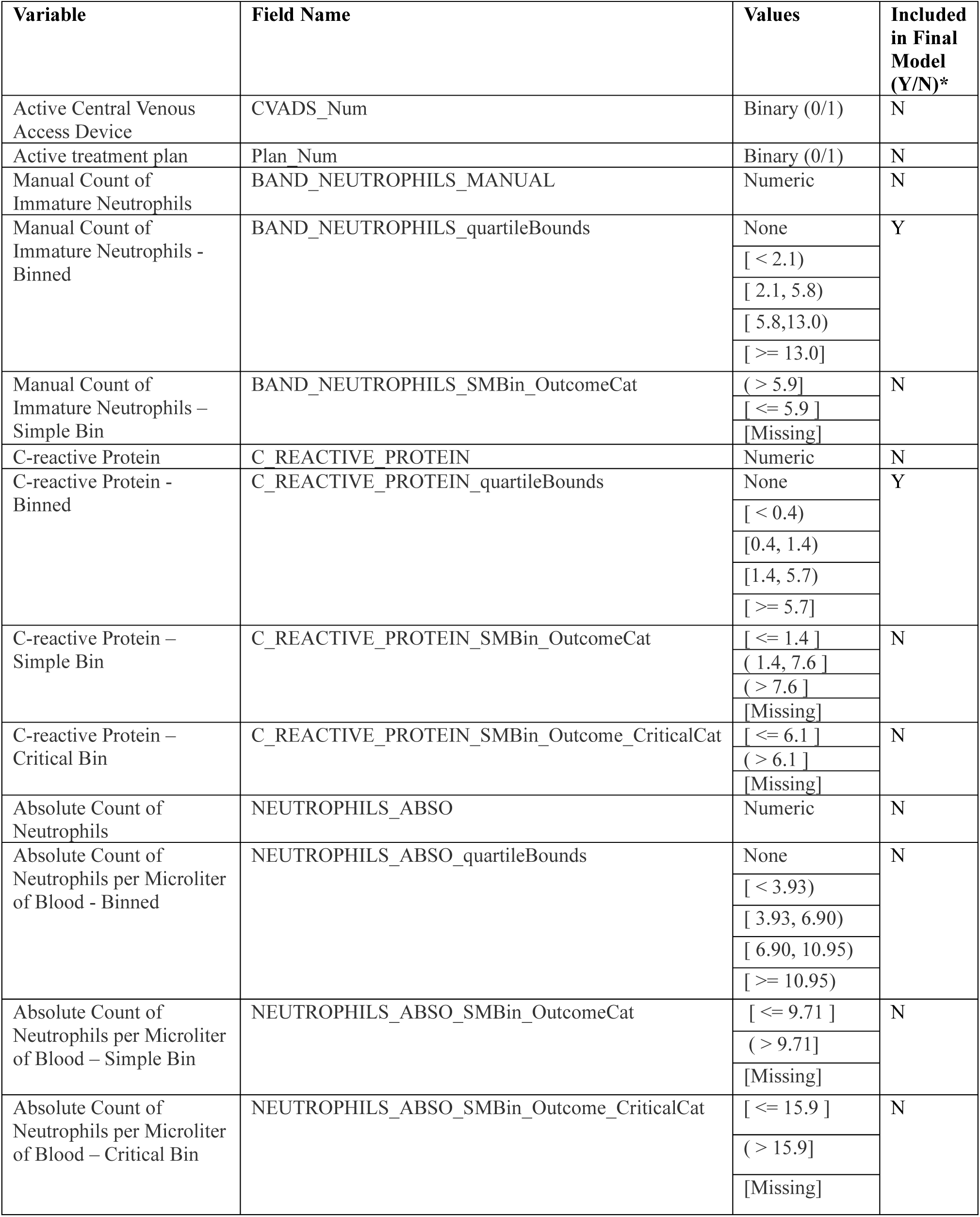
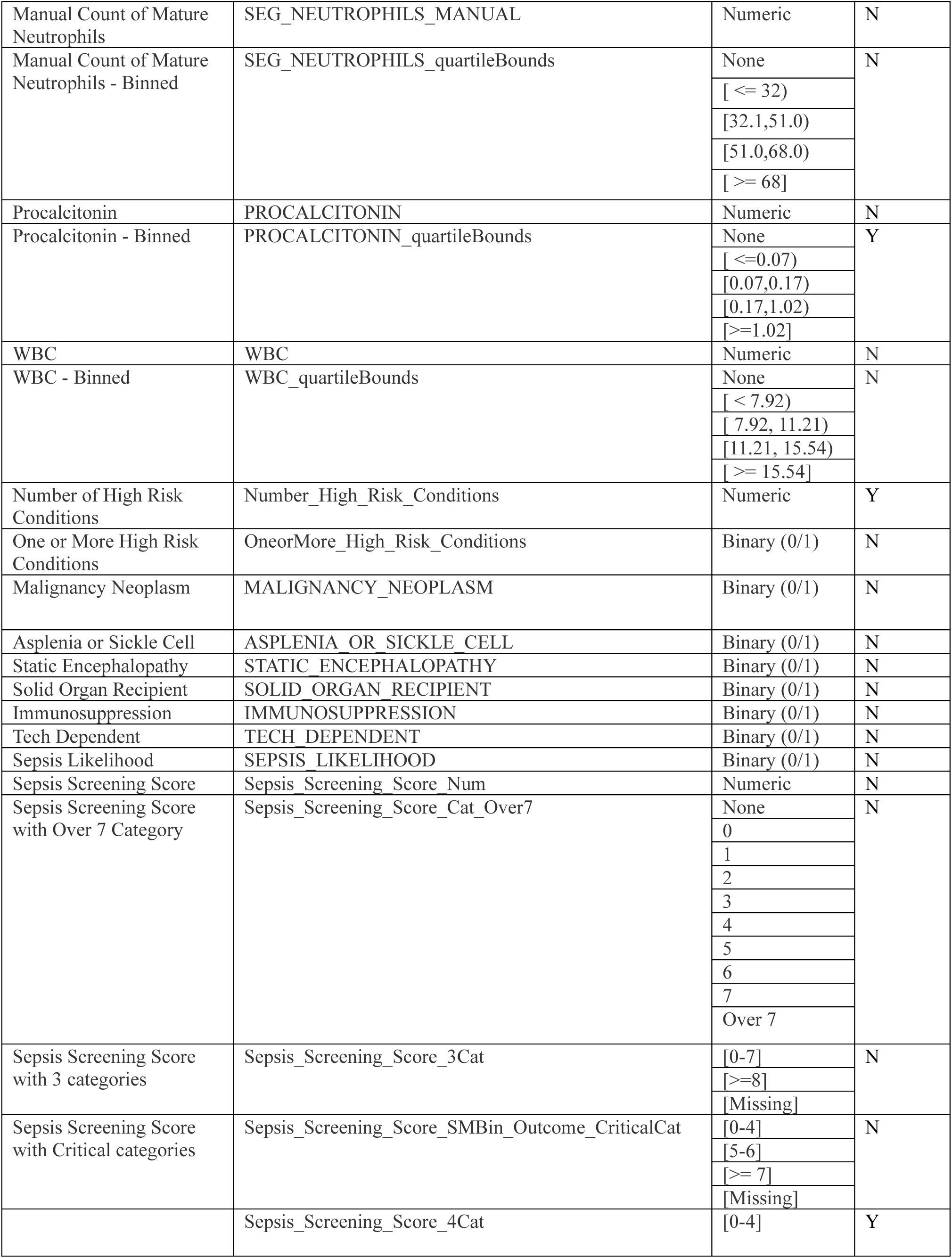
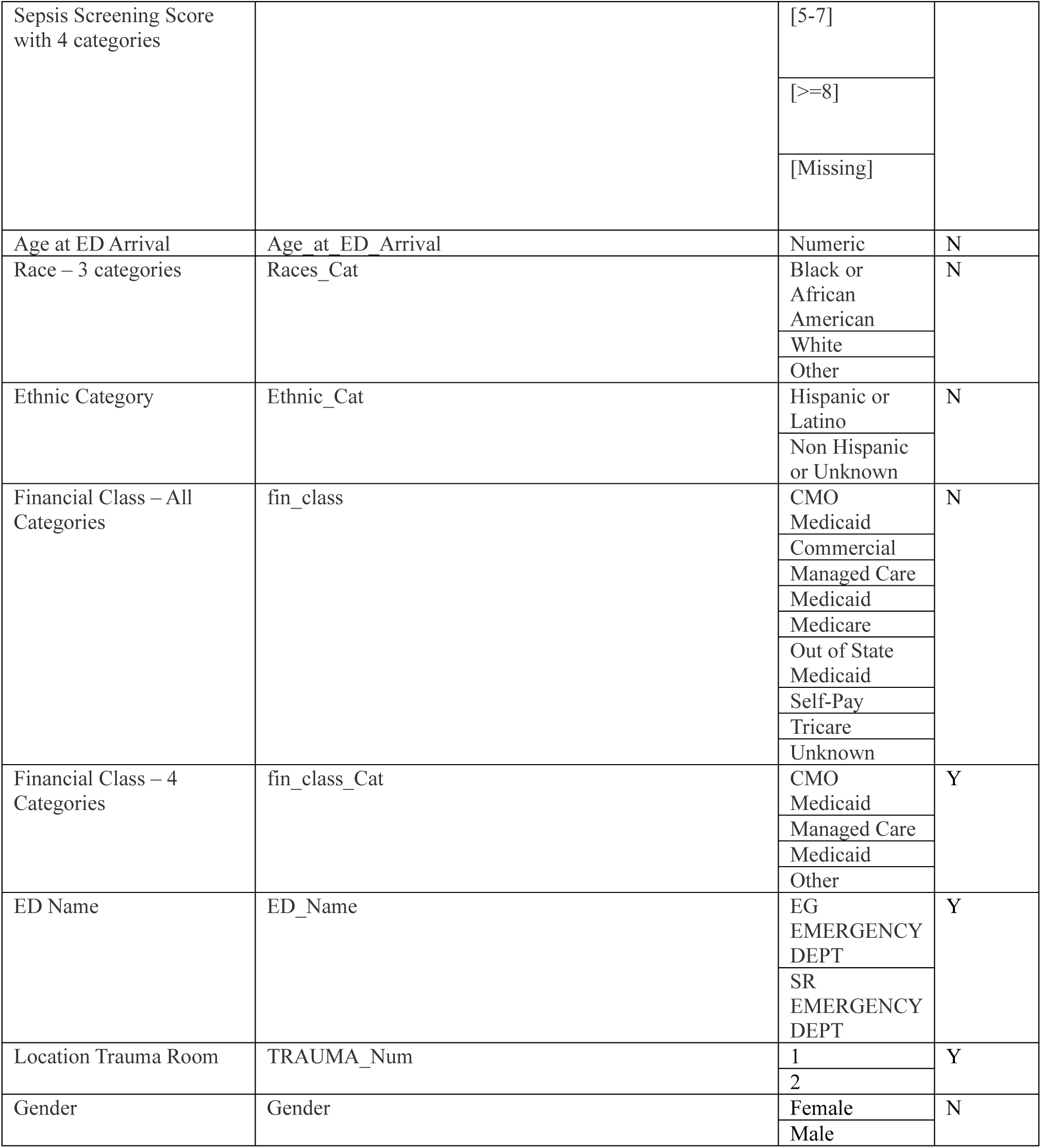
All Model Predictor Variables and Criteria Considered for Model Inclusion.

**Supplemental Table 2:** Sensitivity by Demographics.

| Variable | TP + FN | FN | TP | P value |
| --- | --- | --- | --- | --- |
| **N** | N=34 | N=5 | N=29 |  |
| **Gender:** |  |  |  | 0.445 |
| Female | 19 (55.9%) | 2 (10.5%) | 17 (89.5%) |  |
| Male | 15 (44.1%) | 3 (20.0%) | 12 (80.0%) |  |
| **Race:** |  |  |  | 0.461 |
| Asian | 5 (14.7%) | 0 (0.00%) | 5 (100%) |  |
| Black or African American | 14 (41.2%) | 3 (21.4%) | 11 (78.6%) |  |
| White | 12 (35.3%) | 1 (8.33%) | 11 (91.7%) |  |
| Other | 3 (8.82%) | 1 (33.3%) | 2 (66.7%) |  |
| **Ethnicity:** |  |  |  | 0.142 |
| Non Hispanic or Latino | 32 (94.1%) | 4 (12.5%) | 28 (87.5%) |  |
| Hispanic or Latino | 0 (0%) | 0 (0%) | 0 (0%) |  |
| Other | 2 (5.88%) | 1 (50.0%) | 1 (50.0%) |  |
| **Financial Class:** |  |  |  | 0.231 |
| CMO Medicaid | 13 (38.2%) | 1 (7.69%) | 12 (92.3%) |  |
| Managed Care | 13 (38.2%) | 3 (23.1%) | 10 (76.9%) |  |
| Medicaid | 6 (17.6%) | 0 (0.00%) | 6 (100%) |  |
| Other | 2 (5.88%) | 1 (50.0%) | 1 (50.0%) |  |
| **Age at ED Arrival** | 3.54 [0.17;6.94] | 10.3 [1.75;16.2] | 3.17 [0.17;6.33] | 0.180 |

TP – True Positive, FN – False Negative; Sensitivity for each group is calculated as TP / (TP+FN). Distributional differences for categories are tested by chi-squared or exact Fisher test when the expected frequency is less than 5 in a cell, and by Kruskal-Wallis test for Age at ED Arrival.

**Supplemental Table 3:** Positive Predictive Value (PPV) by Demographics.

| Variable | TP + FP | FP | TP | P value |
| --- | --- | --- | --- | --- |
| **N:** | N = 434 | N = 405 | N = 29 |  |
| **Gender:** |  |  |  | 0.091 |
| Female | 189 (43.5%) | 172 (91.0%) | 17 (8.99%) |  |
| Male | 245 (56.5%) | 233 (95.1%) | 12 (4.90%) |  |
| **Race:** |  |  |  | 0.111 |
| Asian | 25 (5.76%) | 20 (80.0%) | 5 (20.0%) |  |
| Black or African American | 173 (39.9%) | 162 (93.6%) | 11 (6.36%) |  |
| White | 201 (46.3%) | 190 (94.5%) | 11 (5.47%) |  |
| Other | 35 (8.1%) | 33 (94.3%) | 2 (5.7%) |  |
| **Ethnicity:** |  |  |  | 0.084 |
| Hispanic or Latino | 62 (14.3%) | 61 (98.4%) | 1 (1.61%) |  |
| Non-Hispanic or Latino | 372 (85.7%) | 344 (92.5%) | 28 (7.53%) |  |
| **Financial Class:** |  |  |  | 0.794 |
| CMO Medicaid | 186 (44.3%) | 174 (93.5%) | 12 (6.45%) |  |
| Managed Care | 142 (33.8%) | 132 (93.0%) | 10 (7.04%) |  |
| Medicaid | 77 (18.3%) | 71 (92.2%) | 6 (7.79%) |  |
| Tricare | 9 (2.14%) | 9 (100.0%) | 0 (0.00%) |  |
| Other | 6 (1.43%) | 6 (100.0%) | 0 (0.00%) |  |
| **Age at ED Arrival:** | 4.00 [0.83,10.8] | 4.00 [0.92,10.8] | 3.17 [0.17,6.33] | 0.235 |

TP – True Positive, FP – False Positive; Positive Predictive Value (PPV) for each group is calculated as TP / (TP+FP). Distributional differences for categories are tested by chi-squared or exact Fisher test when the expected frequency is less than 5 in a cell, and by Kruskal-Wallis test for Age at ED Arrival.

**Supplemental Figure 1:**
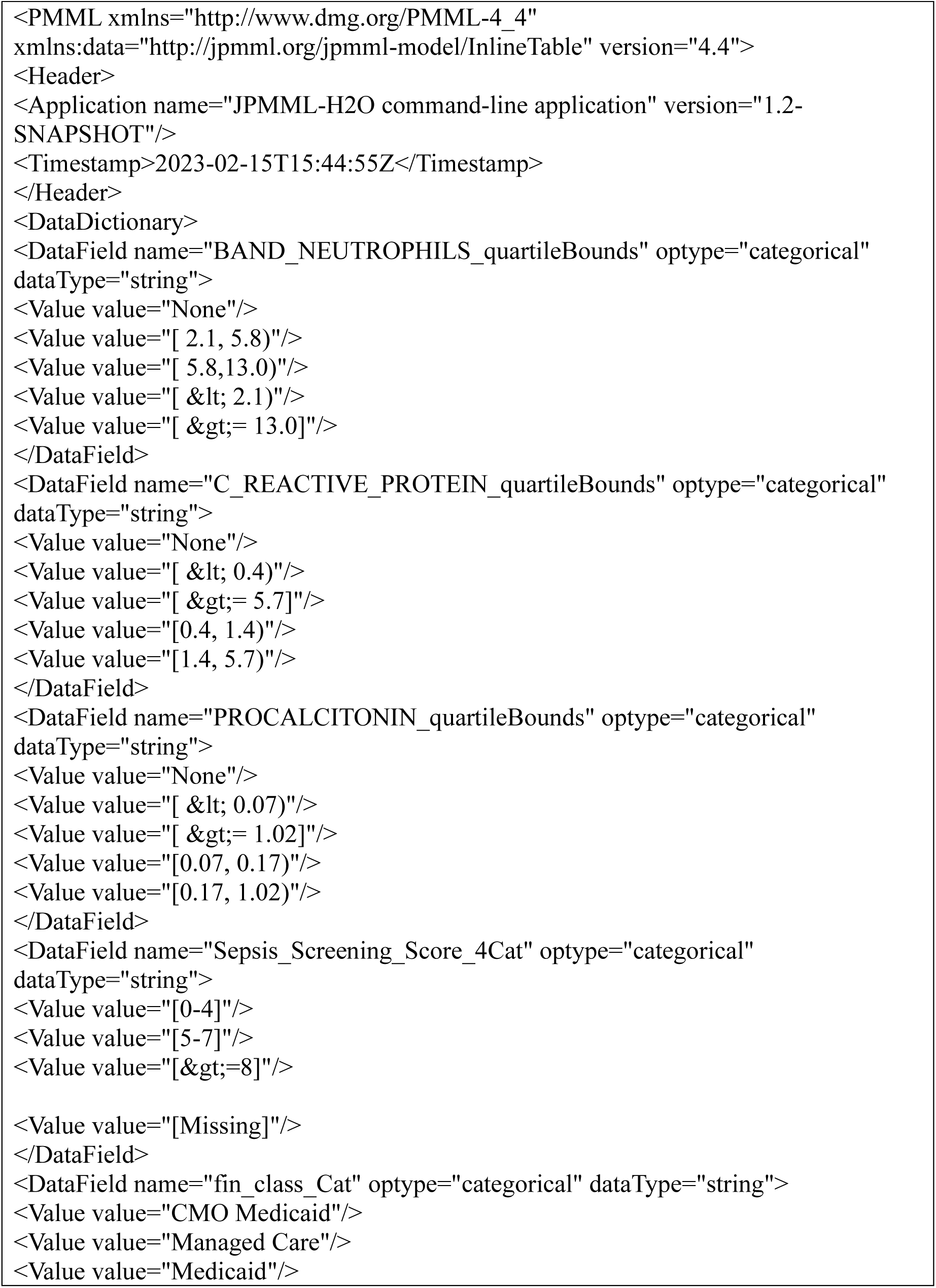
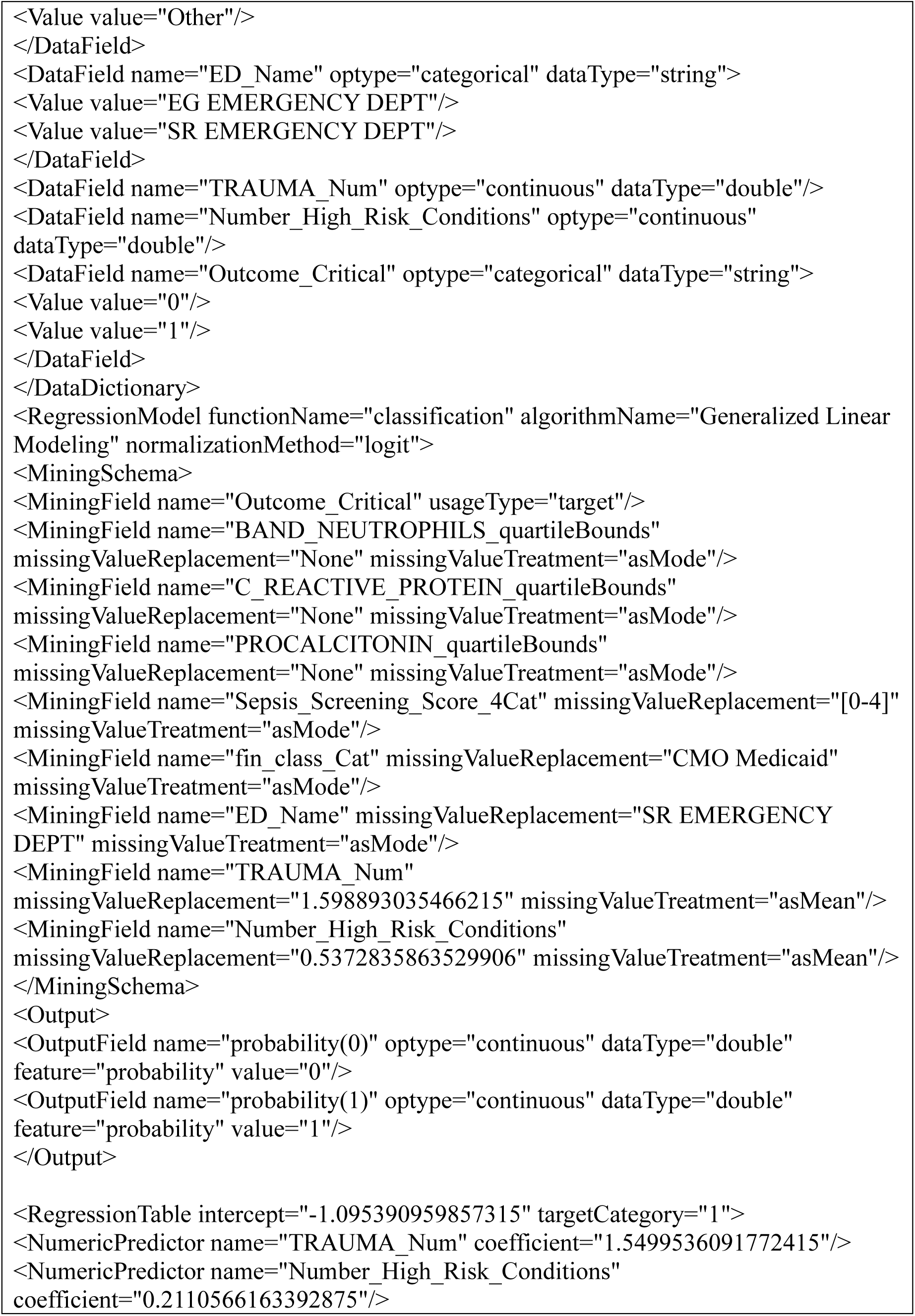
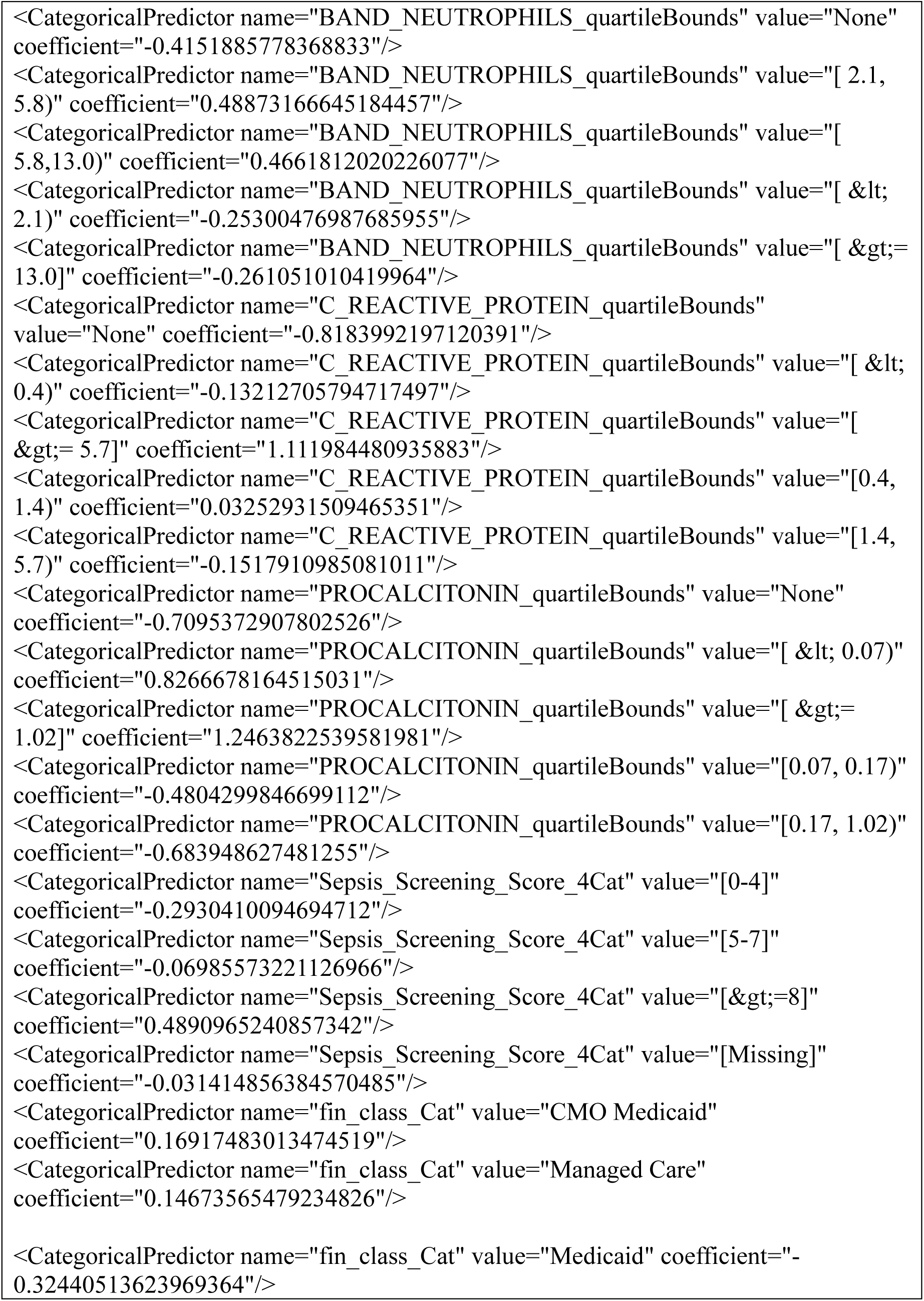
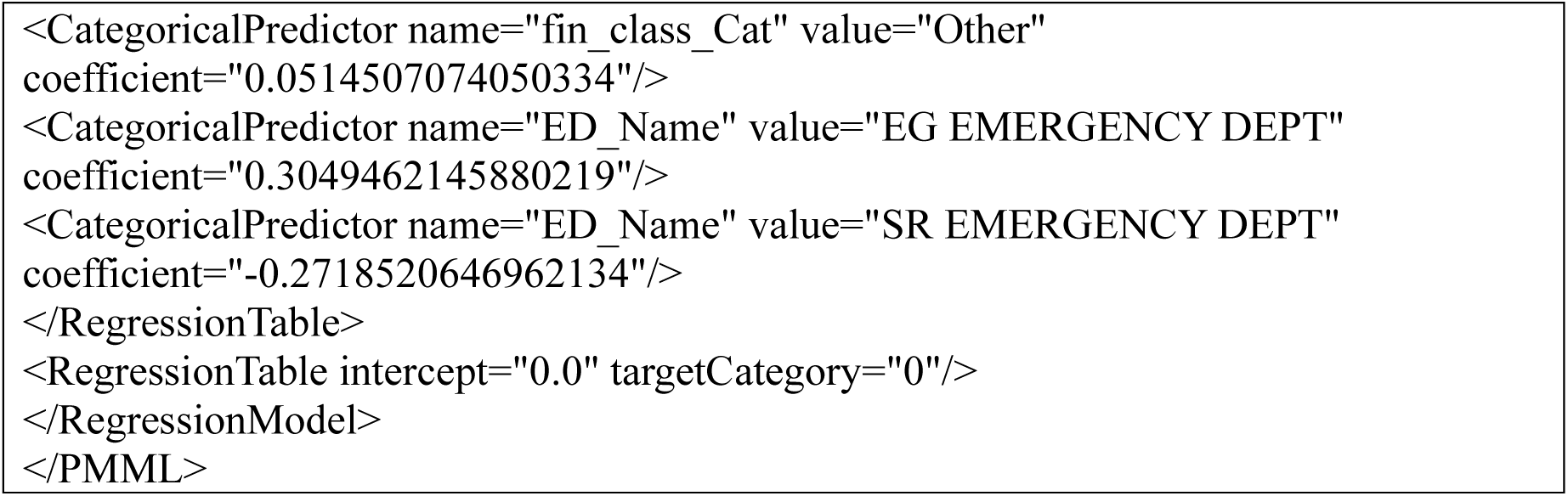
Predictive Model Markup Language (PMML) Code for the SEPD Model

## Notes

**Funding / Support**: This work was supported by the Agency for Healthcare Research and Quality (AHRQ) and the National Center for Advancing Translational Sciences (NCATS) of the National Institutes of Health (NIH) grant numbers 5R03HS029417-02 (SK, EWO, MVM), UL1TR002378 (MVM) and KL2TR002381 (MVM).

### Competing Interest Statement

All authors have completed the ICMJE uniform disclosure form at www.icmje.org/coi_disclosure.pdf and declare: : no support from any organization for the submitted work; EO and NM are co-founders and have equity in Phrase Health a clinical decision support analytics company. They are Investigators on an R42 grant with Phrase Health from the National Library of Medicine (NLM) and National
Center for Advancing Translational Science (NCATS). Both of them receive salary support from the NLM and NCATS, but no direct revenue from Phrase Health.

### Funding Statement

Supported in part by the National Center for Advancing Translational Sciences of the
National Institutes of Health under Award Number UL1TR002378 and KL2TR002381 (MVM).

### Author Declarations

The protocol for this retrospective analysis of model performance was reviewed and deemed not human subjects research by the Children's Healthcare of Atlanta Institutional Review Board.

